# Patient safety culture, teamwork, and observed perioperative safety compliance in a high-volume surgical unit

**DOI:** 10.64898/2026.08.31.26361796

**Authors:** Venus Mandala Sahputri, Agiona Angeline, Erwin Tenggono

## Abstract

Perioperative safety checklists standardize critical actions, but reliable completion depends on the surrounding work system and team behavior. We conducted a prospective observational analytic study from April to May 2026 in the central surgical unit of a high-volume public teaching referral hospital in Indonesia to examine whether patient safety culture and teamwork were associated with directly observed perioperative safety compliance and whether teamwork mediated the culture-compliance relationship. Patient safety culture was measured with the Hospital Survey on Patient Safety Culture 2.0, teamwork with a 35-item TeamSTEPPS Teamwork Perceptions Questionnaire research adaptation, and compliance by direct role-based observation using a 45-item checklist derived from the AORN Comprehensive Surgical Checklist. Eighty of 92 recruited professionals contributed 240 person-operation observations across 50 operations. Overall compliance was 74.75%, with sign-out lowest at 70.68%. Patient safety culture was associated with teamwork (β = 0.590; 95% CI 0.510-0.770) and directly with compliance (β = 0.407; 95% CI 0.187-0.712). The teamwork-compliance coefficient was positive (β = 0.285; p = 0.046), but the prespecified percentile 95% CI included zero (−0.045 to 0.517). The indirect effect through teamwork was not supported (β = 0.168; p = 0.079). These findings support a system-level interpretation of perioperative safety and identify learning-oriented responses to error, situation monitoring, and sign-out fidelity as measurable targets for future improvement efforts.

**Author summary:** Surgical safety checklists are intended to make critical safety actions reliable, but their effectiveness depends on how teams and organizations work. We studied 80 perioperative professionals in a high-volume Indonesian teaching hospital and linked their perceptions of patient safety culture and teamwork with directly observed compliance across 240 person-operation observations. Stronger safety culture was associated with better teamwork and higher observed compliance. Teamwork itself showed a positive but statistically inconsistent association with compliance and did not explain the full culture-compliance relationship. The findings suggest that quality improvement should combine learning-oriented responses to error, active situation monitoring, and stronger sign-out execution rather than relying on teamwork training alone.

## Introduction

Reliable completion of perioperative safety checks is a process-quality issue, not simply a documentation task. Surgical checklists standardize critical actions and team communication, but their presence does not ensure that checks occur completely, at the right time, or with the appropriate participants [1–3]. Patient safety culture and teamwork are two work-system conditions that may shape this reliability. Operating-room research identifies nonpunitive response to error, communication openness, staffing, handoffs, management support, role clarity, psychological safety, and team stability as recurring determinants of safety performance [4–11].

However, much of this evidence is based on self-report measures. Less is known about how perceived safety culture and teamwork relate to directly observed perioperative behavior, or whether teamwork explains the pathway from organizational culture to observed compliance. Systems Engineering Initiative for Patient Safety (SEIPS) 3.0 provides a useful framework because it links organizational work-system conditions, collaborative professional work, and process performance across the patient journey [12].

We therefore examined whether patient safety culture was associated with teamwork and directly observed perioperative safety compliance, tested teamwork as a statistical mediator, and identified the safety-culture and teamwork dimensions most relevant to the higher-order constructs. The practical aim was to identify measurable targets that quality leaders could use when designing future perioperative improvement efforts. These aims were achieved through direct observation combined with hierarchical structural modeling.

## Materials and methods

### Design and setting

We conducted a prospective observational analytic study from April to May 2026 in the central surgical unit of RSUD Dr. Moewardi, Surakarta, Indonesia, a Class A public teaching referral hospital. The unit comprised 12 operating rooms and routinely managed approximately 32 to 51 surgical procedures per day. Questionnaires were completed at the beginning of the study period, followed by direct perioperative observation from April 13 to May 8, 2026. The individual healthcare professional was the unit of analysis; repeated operation-level observations were aggregated to one record per participant. Reporting followed the STROBE statement (S2 Checklist).

### Participants and sampling

Participants were recruited from April 13 to May 8, 2026. Written informed consent was obtained from all participants before questionnaire completion and direct perioperative observation. The source population comprised 140 healthcare professionals actively involved in perioperative care, including anesthesiologists, surgeons/operators, physicians/residents, unit coordinators or head nurses, anesthesia nurses/assistants, scrub nurses, and circulating nurses. Proportionate stratified random sampling by profession was used. A priori planning for a two-predictor regression framework, assuming f^2^ = 0.15, α = 0.05, and 80% power, indicated a minimum of approximately 68 participants; the operational target was at least 80 complete cases. Participants were included in the main analysis when they had a usable questionnaire, at least three observed operations, and at least 10 applicable checklist opportunities.

### Measures and data quality

Patient safety culture was assessed using the Indonesian wording of the Hospital Survey on Patient Safety Culture 2.0 (HSOPSC 2.0), which comprises 10 dimensions and was informed by a published Indonesian cross-cultural adaptation [13].

Teamwork was assessed using a 35-item Indonesian research adaptation of the TeamSTEPPS Teamwork Perceptions Questionnaire (T-TPQ), covering Team Structure, Leadership, Situation Monitoring, Mutual Support, and Communication [14,15].

Perioperative safety compliance was measured by direct role-based observation using a 45-item checklist developed from the AORN Comprehensive Surgical Checklist and adapted to the study workflow [16]. The checklist included 13 preprocedure check-in, nine sign-in, 14 time-out, and nine sign-out actions. Each applicable opportunity was scored 1 when the action was completed at the appropriate time with the required participant(s), and 0 when omitted, incomplete, late, or only documented. Not-applicable opportunities were excluded from the numerator and denominator. Phase-specific compliance and overall compliance (KPT_ALL) were calculated as compliant actions divided by applicable opportunities × 100. Before the main study, readability, face validity, and cognitive review were conducted with eight healthcare professionals with characteristics similar to the target population. A pilot test with 15 professionals assessed internal consistency, and two observers independently assessed interobserver reliability across 10 operations. The T-TPQ translation and AORN-derived observation tool were research adaptations; no expert-panel content-validity study was conducted. Anonymous participant codes, reverse-coding checks, missing-value review, tracking of applicable opportunities, and participant-level aggregation were used to strengthen data quality.

### Statistical analysis

Descriptive statistics were reported as frequencies and percentages or means ± SD, as appropriate. Partial least squares structural equation modeling (PLS-SEM) was conducted in SmartPLS 4 because the analysis combined causal-predictive objectives with reflective-formative hierarchical component models [17,18]. No custom statistical code was used. No imputation was performed; participants with incomplete questionnaires or insufficient prespecified observation opportunities were excluded before the main analysis.

A disjoint two-stage approach was used. Stage 1 evaluated lower-order components using outer loadings, rho_A, composite reliability, average variance extracted (AVE), and heterotrait-monotrait ratios (HTMT). Stage 2 used lower-order component scores to form higher-order patient safety culture and teamwork constructs; KPT_ALL was modeled as a single-indicator endogenous construct. Collinearity was assessed using variance inflation factors, and structural evaluation included path coefficients, R^2^, f^2^, and the specific indirect effect. Sample adequacy was considered against the a priori calculation and contemporary PLS-SEM guidance [19]. Full measurement and predictive results are reported in S1 Appendix.

Final bootstrapping used 10,000 subsamples, two-tailed tests, and 95% percentile confidence intervals (CIs). Inferential support was prespecified as a percentile bootstrap CI that excluded zero; exact p values were reported as complementary statistics. Predictive relevance was assessed with PLSpredict and the cross-validated predictive ability test (CVPAT) [20,21].

### Ethics statement

The Research Ethics Committee (KEP FEB), Faculty of Economics and Business, Universitas Pelita Harapan approved the study on March 24, 2026 (No. 014/MARS/EC/III/2026). The study was conducted in accordance with the principles of the Declaration of Helsinki. Written informed consent for questionnaire completion and direct perioperative observation was obtained from all participants before participation. Participant data were coded and reported in aggregate.

## Results

### Participants and observations

Ninety-two professionals were recruited and 80 (86.96%) met the criteria for the main analysis. Twelve were excluded because they declined observation (n = 4), had incomplete questionnaires (n = 3), had fewer than three observed operations (n = 3), or had fewer than 10 applicable opportunities (n = 2). The 80 participants contributed 240 person-operation observations from 50 unique operations; 42 operations were elective and eight were emergency procedures. Mean operation duration was 142.20 minutes (range 45-240 minutes). Participant characteristics are shown in Table 1.

**Table 1.** Participant characteristics (n = 80).

| Characteristic | n (%) or mean ± SD | Range |
| --- | --- | --- |
| Scrub nurse | 18 (22.50%) | - |
| Circulating nurse | 18 (22.50%) | - |
| Anesthesia nurse/assistant | 12 (15.00%) | - |
| Surgeon/operator | 10 (12.50%) | - |
| Physician/resident | 10 (12.50%) | - |
| Anesthesiologist | 8 (10.00%) | - |
| Coordinator/manager/head nurse | 4 (5.00%) | - |
| Hospital tenure, years | 10.22 ± 5.05 | 1.58-19.33 |
| Central surgical unit tenure, years | 6.23 ± 3.88 | 1.17-15.83 |
| Working hours/week | 46.10 ± 5.10 | 38-59 |
| Operations attended/week | 10.43 ± 3.22 | 3-17 |
| Patient-safety training | 53 (66.25%) | - |
| Surgical checklist/AORN training | 53 (66.25%) | - |
| SBAR communication training | 43 (53.75%) | - |
| TeamSTEPPS/teamwork training | 35 (43.75%) | - |

Abbreviations: AORN, Association of periOperative Registered Nurses; SBAR, Situation-Background-Assessment-Recommendation; TeamSTEPPS, Team Strategies and Tools to Enhance Performance and Patient Safety.

### Safety performance and measurement quality

Mean patient safety culture was 3.20 ± 0.70 on a 1-to-5 scale and mean teamwork perception was 3.36 ± 0.77. Overall perioperative safety compliance was 74.75 ± 12.17%. Compliance was highest during preprocedure check-in (77.11%) and lowest during sign-out (70.68%) (Fig 1; Table 2).

**Table 2.** Perioperative safety performance and structural model findings.

| Measure or path | Estimate | Precision/statistic | Interpretation |
| --- | --- | --- | --- |
| Patient safety culture (1-5) | Mean $3.20 \pm 0.70$ | Range 1.59-4.53 | - |
| Teamwork perception (1-5) | Mean $3.36 \pm 0.77$ | Range 1.37-4.86 | - |
| Preprocedure check-in | $77.11\% \pm 15.09$ | Range 41.67%-100% | - |
| Sign-in | $75.62\% \pm 16.39$ | Range 16.67%-100% | - |
| Time-out | $75.06\% \pm 16.14$ | Range 33.33%-100% | - |
| Sign-out | $70.68\% \pm 15.21$ | Range 42.11%-100% | Lowest phase compliance |
| Overall compliance (KPT_ALL) | $74.75\% \pm 12.17$ | Range 42.11%-100% | - |
| Safety culture → Teamwork | $\beta = 0.590$ | 95% CI [0.510, 0.770]; $p < 0.001$ | Supported; $f^2 = 0.533$ |
| Safety culture → Compliance | $\beta = 0.407$ | 95% CI [0.187, 0.712]; $p = 0.003$ | Supported; $f^2 = 0.175$ |
| Teamwork → Compliance | $\beta = 0.285$ | 95% CI [-0.045, 0.517]; $p = 0.046$ | Not consistently supported; $f^2 = 0.086$ |
| Safety culture → Teamwork → Compliance | $\beta = 0.168$ | 95% CI [-0.031, 0.349]; $p = 0.079$ | Indirect effect not supported |

Interpretive support was prespecified as a 95% percentile bootstrap confidence interval (CI) that excluded zero. $R^2 = 0.348$ for teamwork and 0.383 for perioperative compliance.

**Fig 1.**
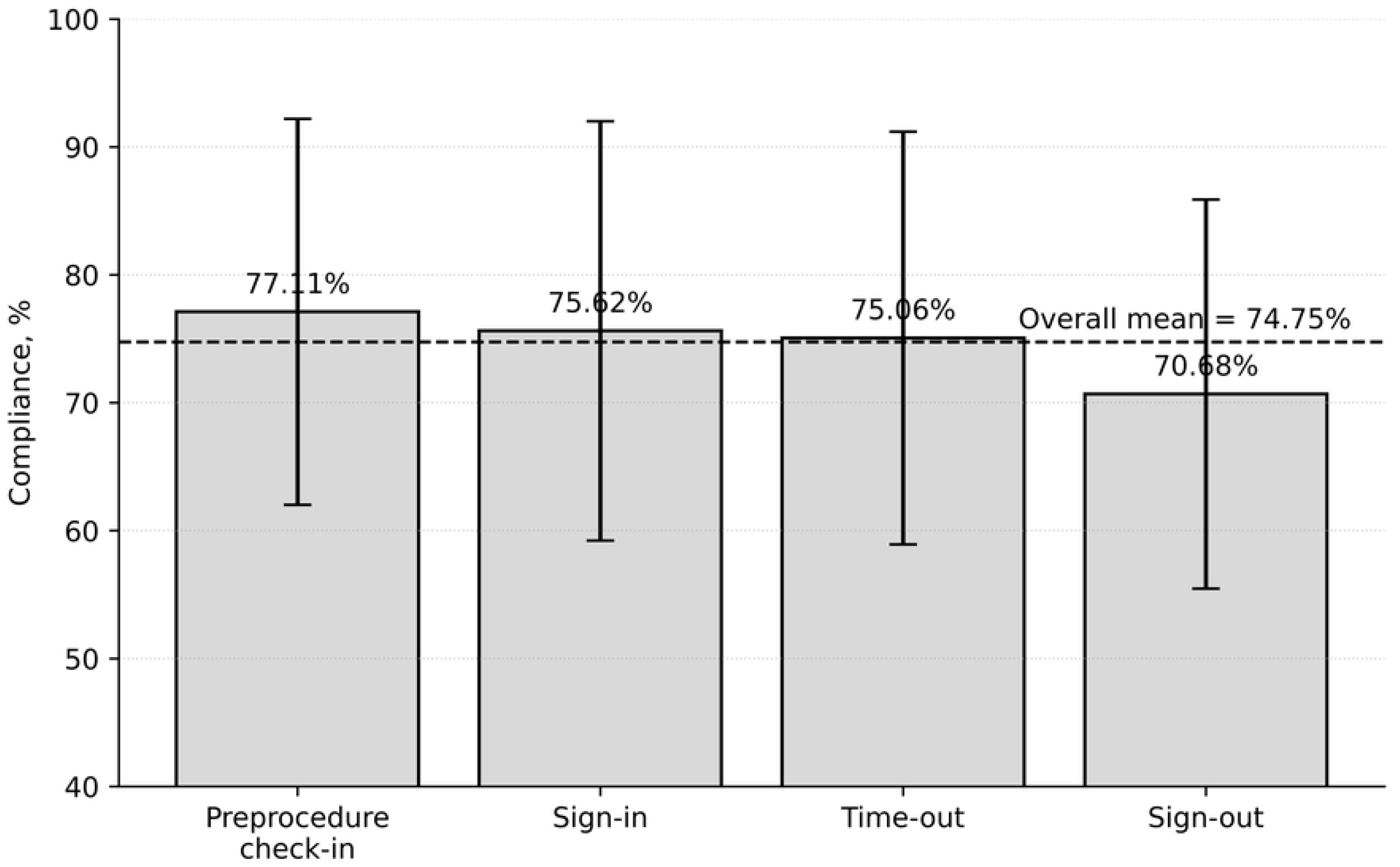
Mean perioperative safety compliance by phase. Error bars indicate SD. The dashed line marks overall mean compliance (74.75%). Sign-out had the lowest mean compliance (70.68%).

At least 92% of item-level readability and face-validity ratings scored 3 or 4 on a four-point scale. Pilot-test Cronbach α was 0.882 for HSOPSC overall and 0.906 for T-TPQ overall. Across 395 applicable paired observer ratings, agreement was 95.19% and Cohen κ was 0.876. The stage-1 measurement model met reliability, convergent-validity, and discriminant-validity criteria; 66 of 67 indicator loadings were at least 0.708, and TTPQ24 (0.680) was retained because construct reliability and AVE remained acceptable. All stage-2 outer variance inflation factors were below 3. Response to Error had the largest supported patient-safety-culture weight (0.525; p = 0.011), and Situation Monitoring had the largest supported teamwork weight (0.583; p = 0.018). Detailed measurement results are provided in S1 Appendix.

### Structural associations and prediction

Patient safety culture was positively associated with teamwork (β = 0.590; 95% CI 0.510-0.770), explaining 34.8% of teamwork variance, and directly with perioperative compliance (β = 0.407; 95% CI 0.187-0.712). The teamwork-compliance coefficient was positive (β = 0.285; p = 0.046), but its prespecified percentile 95% CI included zero (−0.045 to 0.517); this pathway was therefore not considered consistently supported. The indirect effect of safety culture on compliance through teamwork was not supported (β = 0.168; p = 0.079; 95% CI −0.031 to 0.349). Safety culture and teamwork jointly explained 38.3% of compliance variance (Table 2; Fig 2). For KPT_ALL, Q^2^predict was 0.194. PLS-SEM produced slightly lower prediction error than the linear benchmark (RMSE 11.03 vs 11.23; MAE 8.31 vs 8.35), but CVPAT did not show a significant predictive-loss advantage for KPT (p = 0.434) or for the model overall (p = 0.389).

**Fig 2.**
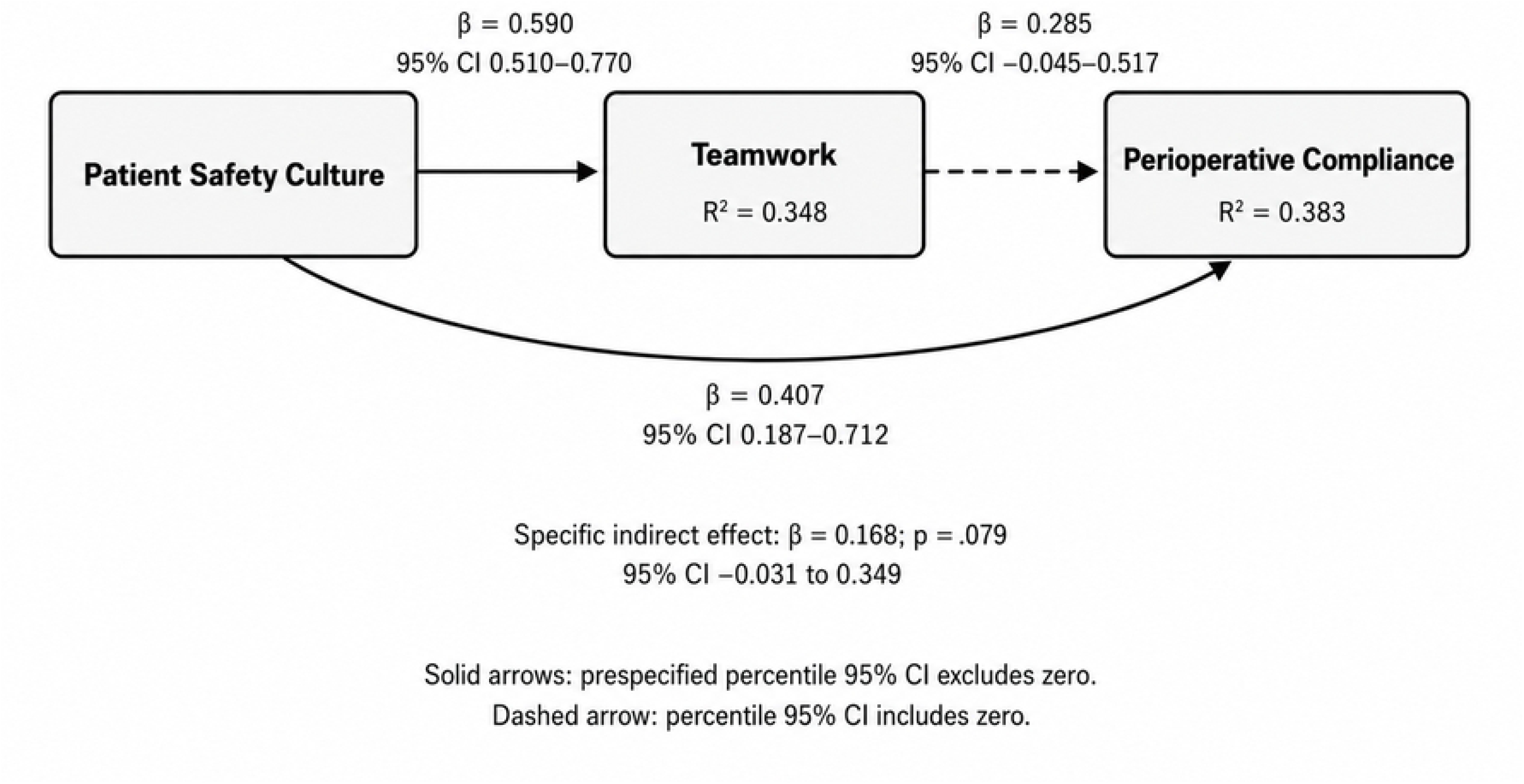
Final structural model. β values are original-sample path coefficients and R^2^ values are shown for endogenous constructs. The teamwork-to-compliance path had p = 0.046, but the prespecified 95% percentile bootstrap confidence interval included zero.

## Discussion

Three findings are central. First, patient safety culture was strongly associated with teamwork. Second, safety culture had a direct positive association with directly observed perioperative compliance. Third, teamwork did not statistically mediate the culture-compliance relationship under the prespecified percentile-bootstrap criterion. Together, these findings suggest that reliable perioperative safety performance reflects a broader work system rather than checklist completion as an isolated individual behavior.

The culture-teamwork relationship is consistent with operating-room evidence showing that hierarchy, role clarity, familiarity, resource constraints, communication, team stability, and psychological safety shape collaborative performance [8–11]. The direct culture-compliance association is also relevant because organizational culture can influence process reliability through supervision, audit and feedback, staffing decisions, leadership responsiveness, and the legitimacy assigned to safety procedures. Checklist reimplementation studies and Indonesian evidence similarly point to organizational support, training, audit, workload, and teamwork as conditions that influence implementation fidelity [1,7].

The teamwork-compliance finding requires cautious interpretation. Although the coefficient was positive and p = 0.046, the prespecified percentile CI crossed zero. The pathway therefore should not be described as robustly supported. This does not imply that teamwork is unimportant: direct observational research links team checklist performance with clinical outcomes, and teamwork remains a recognized determinant of operating-room performance [11,22]. Rather, perceived teamwork may explain only part of the variation in directly observed behavior, and the use of different measurement sources may reduce common-method inflation.

The absence of a significant indirect effect is compatible with a SEIPS 3.0 interpretation in which organizational effects can operate through several parallel pathways rather than a single mediator [12]. Perioperative compliance may simultaneously reflect workload, urgency, resources, leadership, local norms, and implementation routines. A recent systematic review and meta-analysis similarly identified training, a positive work environment, management support, monitoring, and feedback as facilitators of checklist use, while staffing shortages, workload, weak ownership, limited audit, resistance to change, and turnover were barriers [3].

Two component-level findings offer practical targets. Response to Error was the strongest supported patient-safety-culture dimension, reinforcing the importance of learning-oriented review, feedback, and appropriate speaking-up mechanisms [4,5]. Situation Monitoring was the strongest supported teamwork dimension, supporting practices such as cross-monitoring, closed-loop communication, explicit updates when conditions change, and structured debriefing [9,10]. The phase-specific data provide a third target: sign-out had the lowest mean compliance (70.68%). Similar work has identified incomplete end-of-case checklist performance as a recurrent problem [3,23]. Because attention at the end of an operation shifts toward recovery, documentation, room turnover, and preparation for the next case, sign-out fidelity may require specific measurement and redesign rather than assuming that a signed checklist represents complete performance.

### Strengths and limitations

This study combined perception-based instruments with direct role-based observation, repeated operation-level opportunities, interobserver reliability assessment, hierarchical PLS-SEM, mediation analysis, and out-of-sample predictive assessment. However, it was conducted in one high-volume public teaching hospital, which limits generalizability and does not establish causality. Questionnaire responses may be affected by social-desirability bias, and direct observation may have produced a Hawthorne effect. The Indonesian T-TPQ and AORN-derived observation checklist were research adaptations strengthened through cognitive review and reliability testing but were not subjected to expert-panel content validation. Professionals also worked in overlapping teams and operations, creating contextual dependence not fully captured by individual-level PLS-SEM. Case complexity, actual workload, team composition, turnover during procedures, and resource availability were not modeled. Future multisite and multilevel studies should incorporate these factors and prospectively test improvement strategies.

## Conclusion

Patient safety culture was positively associated with teamwork and with directly observed perioperative safety compliance. Teamwork showed a positive but inferentially inconsistent association with compliance and did not mediate the culture-compliance relationship. The findings support a system-level view of perioperative safety in which organizational learning, situation awareness, and checklist fidelity are measurable quality targets. Sign-out warrants particular attention because it had the lowest phase-specific compliance. Future improvement work should prospectively evaluate learning-oriented error-response systems, situation-monitoring practices, and phase-specific sign-out redesign using repeated direct observation and appropriate balancing measures.

## Data Availability

All relevant data are within the manuscript and its Supporting Information files.

## Supporting information

**S1 Appendix**. Extended PLS-SEM measurement and predictive results.

**S2 Checklist**. STROBE checklist for observational studies.

**S3 Data**. De-identified minimal dataset underlying the findings reported in this study.

**S4 Checklist**. PLOS Human Participants Research Checklist.

## Acknowledgments

None.

